# Acceptability, feasibility and appropriateness of intensified health education, SMS/phone tracing and transport reimbursement for uptake of voluntary medical male circumcision in a sexually transmitted infections clinic in Malawi: a mixed methods study

**DOI:** 10.1101/2024.03.27.24304985

**Authors:** Mitch M. Matoga, Evaristar Kudowa, Joachim Chikuni, Mercy Tsidya, Jennifer Tseka, Beatrice Ndalama, Naomi Bonongwe, Esther Mathiya, Edward Jere, Dumbo Yatina, Blessings Kamtambe, Martin Kapito, Mina C. Hosseinipour, Charles S. Chasela, Sara Jewett

## Abstract

**Introduction:** Uptake of voluntary medical male circumcision (VMMC) remains a challenge in many settings. Innovative implementation strategies are required to scale-up VMMC uptake.

**Methodology:** RITe was a multi-faceted intervention comprising transport reimbursement (R), intensified health education (IHE) and SMS/Telephone tracing (Te), which increased the uptake of VMMC among uncircumcised men with sexually transmitted infections (STIs) in Malawi. Using a concurrent exploratory mixed-method approach, we assessed the intervention’s acceptability, feasibility and appropriateness among men with STIs and healthcare workers (HCWs) at Bwaila District Hospital. Participants completed Likert scale surveys and participated in-depth interviews (IDIs) and focus group discussions (FGDs). We calculated percentages of responses to survey items and summarized common themes using thematic analysis. Median scores and interquartile ranges (IQR) were calculated for acceptability, feasibility and appropriateness of each strategy at baseline and end-line and compared using the Wilcoxon signed rank test.

**Results:** A total of 300 surveys, 17 IDIs and 4 FGDs were conducted with men and HCWs between baseline and end-line. The mean age for men in the survey was 29 years (SD ±8) and most were married/cohabiting (59.3%). Mean age for HCWs was 38.5 years (SD ±7), and most were female (59.1%). For acceptability, participants agreed that RITe was welcome, approvable, and likable. Despite participants agreeing that RITe was a good idea, culture and religion influenced appropriateness, particularly at baseline, which improved at end-line for Te and R. For feasibility, HCWs agreed that RITe was easy to implement, but expressed concerns that R (end-line median = 4, IQR: 2, 4) and Te (end-line median = 4, IQR: 4, 4), were resource intensive, hence unsustainable. Interviews corroborated the survey results. Participants reported that IHE provided important information, Te was a good reminder and R was attractive, but they reported barriers to R and Te such as electricity, limited access to phones and distrust in the government.

**Conclusions:** The RITe intervention was acceptable, feasible and appropriate. However, culture/religion and structural barriers affected perceptions of appropriateness and feasibility, respectively. Continued awareness raising on VMMC and addressing setting-specific structural factors are required to overcome barriers that impede demand-creation interventions for VMMC.

**Study registration:** ClinicalTrials.gov identifier: NCT04677374. Registered on December 18, 2020.

## INTRODUCTION

Voluntary medical male circumcision (VMMC) is an evidence-based intervention for HIV prevention. VMMC reduces the risk of female-to-male HIV transmission by 60%[1–3] and protects against several sexually transmitted infections (STIs) [4]. Since 2007, VMMC coverage has increased in 15 high-priority countries in eastern and southern Africa with an estimated 26.8 million boys and men circumcised between 2008 and 2019, resulting in an estimated 340 000 new infections averted by 2019 [5]. In 2016, the Joint United Nations Program on HIV/AIDS (UNAIDS) set a global Fast-Tract target to circumcise an additional 25 million boys and men in high-priority countries by 2020. About 15 million VMMCs were conducted between 2016 and 2020, but this number fell 40% short of the target [5,6].

VMMC uptake has varied across the priority countries. Some of the barriers that have affected uptake of VMMC in the region include culture; religion; fear of infection, bleeding, pain and complications; limited access to VMMC services; costs associated with accessing VMMC; fear of income loss during the recovery period; being attended to by a female provider; and misconceptions/lack of knowledge [7–11]. Other factors have facilitated the uptake of VMMC in the region such as perceived VMMC health benefits, perceived enhanced sexual performance and sexual pleasure among partners of circumcised men, peer/partner pressure, need for sociocultural conformity, and advice from health personnel [7–12].

Malawi adopted VMMC as a HIV prevention strategy in 2012. Despite the rapid scale-up of coverage in recent years, the country was 38% short of its 2,458,727 VMMC target by 2020,[13] and prevalence of circumcision to date (medical and traditional) remains low around 32% [14]. In an attempt to address some of the barriers and to increase VMMC uptake, we implemented a multi-faceted demand-creation strategy intervention consisting of transport **<u>R</u>**eimbursement, **I**ntensified health education and short message service (SMS)/**<u>Te</u>**lephonic tracing (the RITe intervention), which increased VMMC uptake by 100% among uncircumcised men with STIs at Bwaila District Hospital in Lilongwe, Malawi. To better understand the implementation process and to assess the potential for scale-up of our intervention, we evaluated the acceptability, feasibility and appropriateness of the RITe intervention among men with STIs and healthcare workers (HCWs).

## METHODS

Our manuscript followed the Good Reporting for A Mixed Methods Study (GRAMMS) checklist (S1 GRAMMS checklist).

### Study Design and Overview

We conducted a cross-sectional survey using a concurrent exploratory mixed-methods approach nested within a larger implementation trial. We used a mixed-methods approach to have a comprehensive assessment of and to better understand the acceptability, feasibility and appropriateness of the RITe intervention among men and healthcare workers. The primary trial was a pragmatic pre- and post-interventional quasi-experimental study that measured baseline VMMC uptake and prospectively observed uncircumcised men with STIs for 30 days after implementation exposure for uptake of circumcision in Lilongwe, Malawi. The study demonstrated a 100% increase in VMMC uptake between the pre- and post-intervention periods [15]. The intervention was rolled-out in a sequential and incremental manner called implementation blocks. Implementation block 1 was **I**ntensified health education, block 2 was **I**ntensified health education and SMS/telephonic **<u>T</u>**racing and Block 3 was **I**ntensified health education, SMS/telephonic **<u>T</u>**racing and transport **<u>Re</u>**imbursement (RITe).

Intensified health education (IHE) was comprehensive regular group health education (provided at least four sessions a day) tailored to men with STIs followed by one-on-one counseling by a trained mobilizer for interested men. An education guide was developed using expert opinion and literature with guidance from the Malawi VMMC communication strategy [13]. A trained VMMC mobilizer provided health education sessions with support from trained satisfied male and female clients (VMMC champions). All uncircumcised men who presented to the STI clinic during the study period were sensitized on circumcision. SMS/telephonic (cell phone) tracing (“Te”) involved short message reminders to uncircumcised men sent on day 1, 3 and 7 after a STI clinic visit, followed by a phone call on day 14 for those who remained uncircumcised. Lastly, transport reimbursement (“R”) was a compensation of US $10 provided to men who were confirmed to have received circumcision during the study period. Men who expressed interest in circumcision during each block were registered by the VMMC mobilizer and followed up for 30 days for uptake of circumcision.

Quantitative data were collected through Likert-scale surveys using the same men at baseline and end-line of each block. The surveys were also administered to HCWs at baseline and end-line.

Qualitative data was collected through focus group discussions (FGDs) with the same men at baseline and end-line, and through in-depth interviews (IDIs) with the same HCWs at baseline and end-line.

### Study Setting

The study was conducted at the Bwaila District Hospital, a public secondary level health facility in Lilongwe, Malawi. The hospital is situated in a busy central town area and is the only facility with a specialized STI clinic in Lilongwe. The STI clinic sees approximately 60-80 patients per day and is staffed by clinicians, nurses, integrated testing services assistants, and counsellors. As standard practice, all patients receive group health education on HIV and STIs and are offered routine opt-out HIV testing, before receiving care for their STI. During the study period, health education was not offered and VMMC was not routinely offered at the clinic despite this being policy.

Free VMMC services are offered by the Ministry of Health as a ‘walk-in’ outpatient service. However, symptomatic STI patients are advised to receive circumcision after completion of treatment and resolution of symptoms. Just before study implementation, VMMC services were suspended in Malawi due to the COVID-19 pandemic for about 6 months. After the suspension was lifted, Bwaila hospital discontinued providing VMMC services as the VMMC clinic was repurposed to aCOVID-19 isolation center, as a result, men interested in circumcision were referred to their nearest facility.

### Recruitment, sampling and sample size

Participants were recruited from 01 February 2021 to 31 August 2021. Recruitment and sampling were largely convenient. Men who participated in the study were recruited by the VMMC mobilizer and VMMC champions during the IHE sessions. All men provided written informed consent to participate in the surveys and FGDs. For the baseline surveys, we recruited 146 uncircumcised men in total. To assess change in perceptions of acceptability and appropriateness following exposure to the intervention strategies, 132 of the 146 men participated in the end-line surveys. End-line surveys were initially restricted to circumcised men only but due to low incidence of circumcision, we included men involved in the baseline survey who were exposed to the intervention regardless of their circumcision status. We recruited 16 uncircumcised men for the two baseline FGDs (8 men in each session) before the intervention roll-out. We also conducted two end-line FGDs (5 men and 6 men in each session) with both uncircumcised and men who received circumcision during the study period to assess perceptions and experiences on the intervention strategies in increasing VMMC uptake following exposure to the intervention.

The investigator and qualitative research assistants (RAs) recruited interested HCWs from within the STI clinic to participate in the surveys and IDIs. There were 22 HCWs eligible to participate at both baseline and end-line. All HCWs were invited to participate. Baseline IDIs and surveys were conducted before study roll-out. End-line IDIs and surveys were conducted towards the end of the implementation period (end-line). We recruited 12 HCWs for the baseline survey of whom10 completed the end-line survey. For the IDIs, 10 HCWs participated in the baseline interviews of whom 7participated in the end-line interviews.

### Data Collection

#### Quantitative

Quantitative and qualitative data were collected at roughly the same time and independently from each other. Before data collection, all HCWs, qualitative RAs, the VMMC mobilizer and champions were trained by the first author on the study protocol and the data collection tools. The VMMC mobilizer and champions were also sensitized on basic information on HIV, STIs and VMMC and refreshed on research ethics and psychosocial counseling.

Following written informed consent, participants completed a 5-point Likert-scale survey on acceptability, feasibility and appropriateness. Acceptability and appropriateness surveys were administered to both men and HCWs while feasibility surveys were administered only to HCWs at end-line, following exposure to the intervention. The surveys were adapted and validated by the investigator from the acceptability of intervention measure (AIM), intervention appropriateness measure (IAM) and feasibility of intervention measure (FIM), which are psychometric measurement tools [16] for acceptability, appropriateness and feasibility, respectively as defined by Proctor et al [17]. The implementation outcomes were defined as follows:

- Acceptability: the perception among men and HCWs that the RITe intervention is (baseline) or was (endline) agreeable, palatable and satisfactory.
- Appropriateness: the perception among men and HCWs that the RITe intervention is (baseline) or was (endline) a good fit or relevant.
- Feasibility: the extent to which RITe intervention could be (baseline) or was (endline) successfully implemented at Bwaila STI clinic.

For acceptability, participants rated each strategy based on the following constructs: like/appeal, approve, willing, welcome and non-invasive to privacy. For appropriateness, each strategy was rated based on its fit with culture, and religion, whether it was a good idea and not embarrassing. Feasibility of strategies was rated based on ease of implementation, possibility to be routine activity, implementable over time (sustainability), and availability of resources for implementation. Participants rated each construct using a 5-point Likert scale depending on their level of agreement or disagreement to the statements as: “Completely Agree”, “Agree”, “Neither”, “Disagree” and “Completely Disagree”. Surveys were self-administered for HCWs and assisted for men using Open Data Kit Collect v1.4.11 on electronic tablets (S2 survey tools). The survey also included demographic data including age, marital status and educational level for men and age, sex, cadre, and number of years of experience for HCWs.

Data were downloaded from the tablets on to a server every day and saved on a Microsoft Excel spreadsheet by a data officer. Data quality control and assurance checks were done weekly by a data officer.

#### Qualitative

FGDs and IDIs were conducted at the UNC Project Bwaila research site building within the Bwaila district hospital. Participants were escorted by the mobilizer and champions to the interview rooms. The FGDs and IDIs were conducted by the investigator and two qualitative RAs, who also took field notes. Both FGDs and IDIs were audio recorded. IDIs lasted around 60-90 minutes while FGDs lasted around 90-120 minutes. All interviews (FGDs and IDIs) were conducted in the local language (Chichewa).

Interview guides were developed by the investigator and a qualitative research expert. The interview guides were developed based on the quantitative survey tools AIM, IAM and FIM to complement the quantitative data. . The interview guides were piloted and revised before implementation. Similar to the surveys, we collected demographic data on age, sex, cadre and years of experience for IDIs and age, marital status, and education for FGDs.

Interview questions included exploring knowledge of VMMC including benefits and risks, barriers, facilitators to uptake of VMMC, myths and misconceptions about VMMC, perceptions/experiences of the RITe intervention strategies on increasing demand for uptake of VMMC, and the feasibility of delivering the RITe strategies in the STI clinic, among other questions (S3 interview guides).

All participants for the interviews and surveys were compensated with a $10-equivalent in Malawi Kwacha. Interview notes were kept under a ‘double lock’ system only accessible to the investigators. Audio recordings were uploaded weekly to an encrypted file on the UNC Project server and deleted from the recorders.

### Analysis

#### Quantitative

Descriptive statistics including means and proportions were used to summarize demographic characteristics of participants. Survey responses were coded as: “Completely Agree=5”, “Agree=4”, “Neither=3”, “Disagree=2” and “Agree=1”. We calculated percentages for each response to the constructs of acceptability, feasibility, and appropriateness surveys. We plotted histograms for each response to the survey constructs to display the distribution of responses. Median scores and interquartile ranges (IQR) were calculated for the constructs and summarized as an average score for acceptability, feasibility and appropriateness at baseline and end-line. The median scores for acceptability and appropriateness were compared between baseline and end-line using the Wilcoxon signed rank test with corresponding p-values. Statistical analysis was conducted using StataCorp, LLC, College Station, TX, USA, version 16.

#### Qualitative

Audio recordings were concurrently transcribed verbatim and translated into English by two bi-lingual qualitative RAs. Field notes from the interview were summarized immediately after the interviews. After the first FGD and 3 IDIs, we immediately reviewed the recordings and transcriptions for completeness and data quality. Moving forward, a random sample of subsequent interviews was regularly reviewed for data quality by the investigator. Analysis was done by the investigator and RAs using the thematic content analysis approach [18]. A coding scheme was developed using a priori codes for the constructs and combined with inductive codes generated during data analysis. Double coding was done on all transcripts by the investigator and RAs. All coding discrepancies were discussed by the investigator and the qualitative RAs, and the last author assisted with resolving coding discrepancies. Codes were then categorized into salient themes and sub-themes. We used NVivo Version 12, QSR International, Burlington, MA, USA, organize data for analysis. Audit trails were maintained for all qualitative data, including audio recordings, transcripts, notes and all coding documents.

### Ethical consideration

This study was approved by the National Health Sciences Research Committee of Malawi (NHSRC#: 19/10/2412), the University of the Witwatersrand Human Research Ethics Committee (HREC#: M200328) and the University of North Carolina at Chapel Hill Ethics Review Board (IRB#: 19-2559). The main trial was registered with the United States National Library of Medicine (NLM) at the National Institutes of Health Clinical Trial Registry (NCT04677374). All participants provided written informed consent before trial participation. Informed consent was administered by a trained data clerk for the surveys and by a qualitative research assistant for the FGDs and IDIs. The data clerk and qualitative research assistants guided the participants on how to sign the informed consent form. Illiterate participants provided a thumbprint as a sign of consent in the presence of an independent literate witness who co-signed the consent form.

## RESULTS

### Quantitative findings

For the surveys among men, 146 uncircumcised men were recruited for the baseline surveys. Among the 146 men, 132 men (49 circumcised and 83 uncircumcised) men participated in the end-line surveys. Mean age at baseline was 29 years (SD ±6) and 28 years (SD ±7) at end-line. Most men were married or cohabiting with a partner (60.3% baseline, 59.9% end-line) and had secondary level education (52.1% baseline, 50.0% end-line). Demographic characteristics were similar between baseline and end-line participants, and between circumcised and uncircumcised men. Among HCWs, we recruited 12 participants for the baseline surveys of whom 10 participated in the end-line surveys. The mean age among HCWs was 38 years (SD ±6) at baseline and 38 years (SD ±8) at end-line. HCWs were mostly female (58.3% baseline, 60.0% end-line), nurses (59.1%) and with about 10 years of experience. The demographic characteristics were similar between HCWs at baseline and end-line (**Table 1**).

**Table 1:** Demographic characteristics of men and healthcare workers who participated in surveys at Bwaila STI Clinic.

| Characteristic | Men - Surveys |  |
| --- | --- | --- |
|  | Baseline N=146 | *End-line N=132 |
| Age (years), mean (±SD) | 29 (6) | 28 (7) |
| Age categories (years), n (%) |  |  |
| 18-29 years | 80 (54.8) | 66 (50.0) |
| 30-39 years | 37 (25.3) | 36 (27.3) |
| $\geq 40$ years | 29 (19.9) | 30 (22.7) |
| Marital status n (%) |  |  |
| Single | 53 (36.3) | 49 (37.1) |
| Married/Cohabiting | 88 (60.3) | 79 (59.9) |
| Divorced | 4 (2.7) | 4 (3.0) |
| Widowed | 1 (0.7) | 0 (0.0) |
| Education n (%) |  |  |
| None | 4 (2.7) | 3 (2.3) |
| Primary | 55 (37.7) | 53 (40.2) |
| Secondary | 76 (52.1) | 66 (50.0) |
| >Secondary | 11 (7.5) | 10 (7.5) |
| HCWs – Survey |  |  |
|  | Baseline N=12 | *End-line N=10 |
| Age (years), mean (±SD) | 38 ($\pm 6$) | 38 ($\pm 8$) |
| Sex n (%) |  |  |
| Male | 5 (41.7) | 4 (40.0) |
| Female | 7 (58.3) | 6 (60.0) |
| Cadre |  |  |
| Nurse | 7 (58.3) | 6 (60.0) |
| ITS counselor | 4 (33.3) | 3 (30.0) |
| Data clerk | 1 (8.3) | 1 (10.0) |
| **Experience (years), Mean (±SD)** | **10 (±5)** | **10 (±4)** |

SD- Standard deviation, HCW- Healthcare workers, FGDs - Focus group discussions, IDI – In-depth interviews.
\* Participants at end-line were a subset of participants at baseline.

### Acceptability, Feasibility and Appropriateness of IHE among men and HCWs

**Figure 1** shows distribution of Likert scale scores for the acceptability, feasibility and appropriateness of intensified health education at baseline and end-line among men and HCWs. For acceptability, most men and HCWs completely agreed or agreed that IHE was welcome, approvable, likable, and appealing both at baseline and end-line (**Figures 1A and 1B**). Average median score was 5 (IQR:5, 5) at both baseline and end-line for men, and 5 (IQR: 4,5) at baseline and end-line for HCWs (**Table 2**). For appropriateness, most men and HCWs completely agreed or agreed that IHE was a good idea and not embarrassing to discuss in public however, some men (approximately 30%) disagreed that IHE was a good fit to their culture and religion at baseline, with more (slightly over half) who disagreed at end-line in contrast to HCWs who rated IHE as a good fit to culture and religion (**Figures 1C and 1D**). Among men, the average median score dropped from 5 (IQR: 3,5) at baseline to 4 (IQR: 2, 5) (p=0.02) at end-line (**Table 2**). Finally, most HCWs completely agreed or agreed to all the constructs of feasibility with an average median score of 4 (IQR:4, 5) (**Table 2, Figure 1E**).

**Figure 1:**
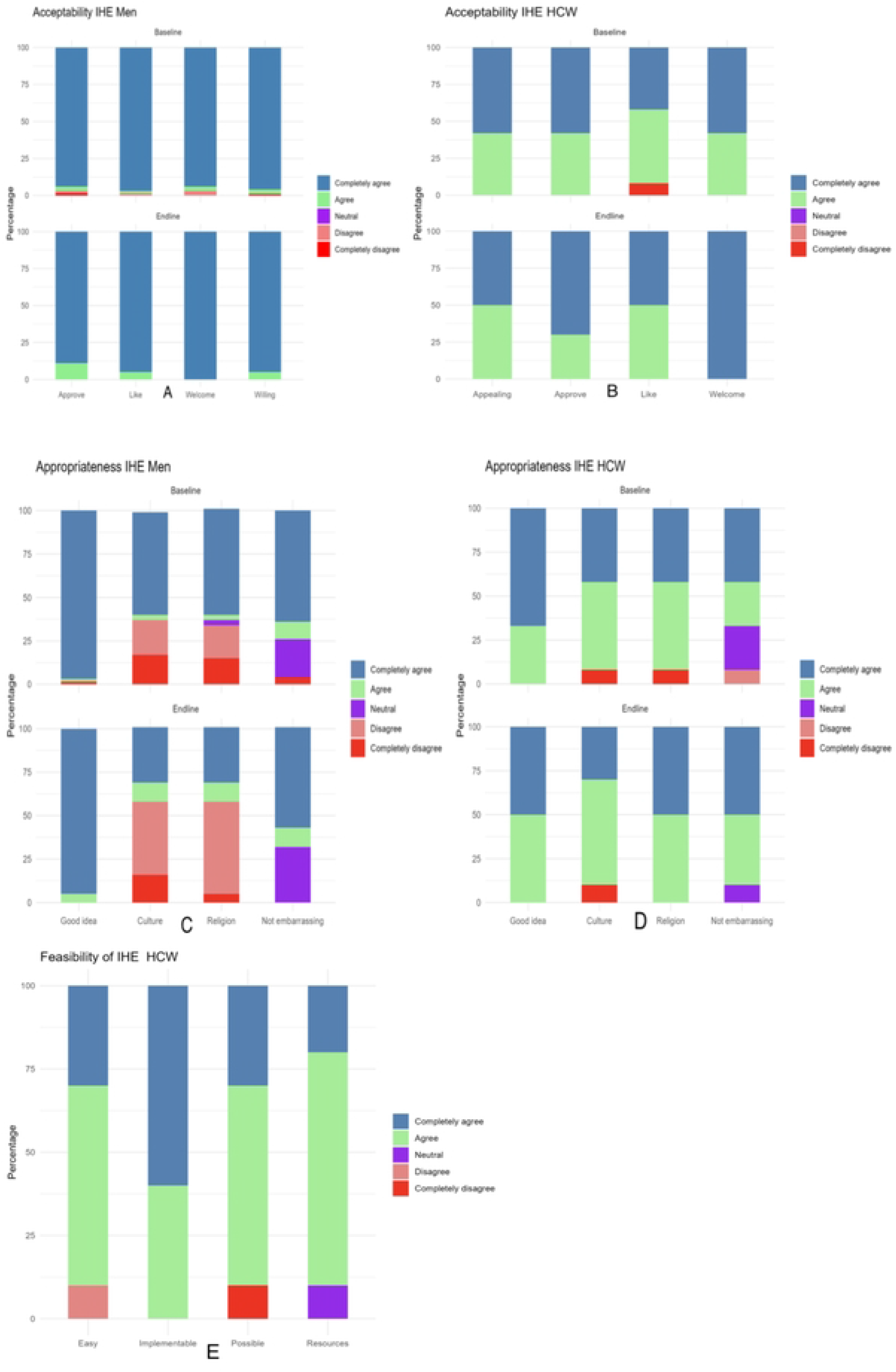
Distribution of Likert scale scores for acceptability, feasibility and appropriateness of intensified health education among men and HCWs at Bwaila STI Clinic at baseline and end-line.

**Table 2:**
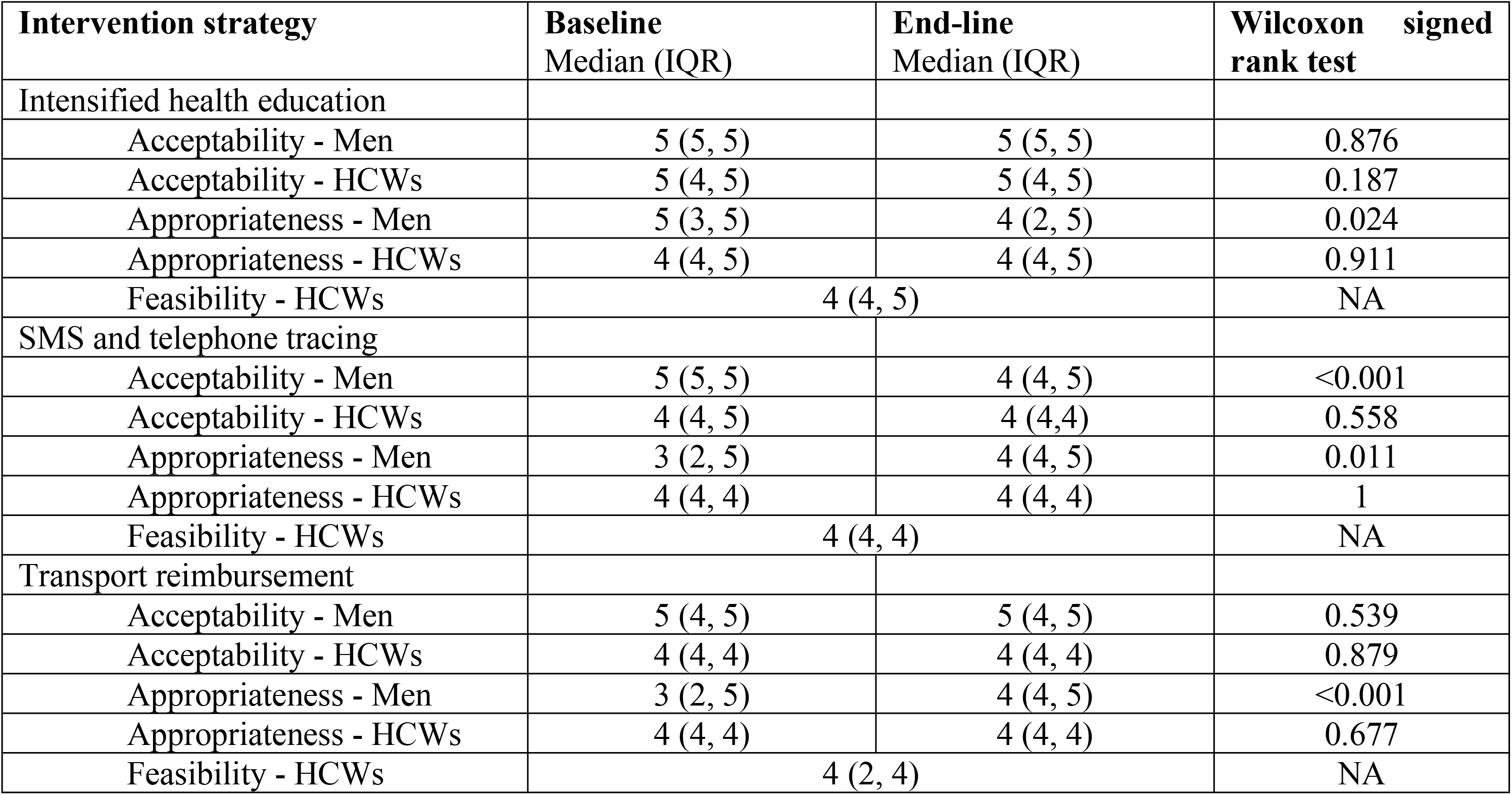
Median scores for acceptability, appropriateness and feasibility at baseline and end-line among men and healthcare workers at Bwaila STI clinic.

### Acceptability, Feasibility and Appropriateness of SMS/telephone tracing among men and HCWs

**Figure 2** shows distribution of Likert scale scores for acceptability, feasibility and appropriateness of SMS/phone call tracing among men and HCWs at baseline and end-line. For acceptability, most men and HCWs completely agreed or agreed that SMS/phone call tracing was welcome, approvable, likable and non-invasive to their privacy at baseline and end-line (**Figures 2A and 2B**). For appropriateness, most men and HCWs completely agreed or agreed that SMS/phone call tracing was a good idea and non-embarrassing **(Figure 2C and 2D)**. However, over half of men at baseline disagreed or completely disagreed that SMS/phone call tracing for VMMC was a good fit to their culture and religion (median = 3, IQR: 2, 5) (**Figure 2C**). Upon exposure to the intervention, this perception improved at end-line (median = 4, IQR: 4, 5) (p=0.01) (**Table 2, Figure 2C**). HCWs generally rated SMS/phone call tracing as a good fit to culture and religion as most completely agreed or agreed with a few that disagreed (less than 15%) at baseline and all fully agreeing at end-line (median = 4, IQR: 4,4) **(Table 2, Figure 2D)**. Overall, SMS/phone call tracing was deemed feasible, but HCWs felt there weren’t adequate resources to implement this strategy as routine practice (median = 4, IQR: 4, 4) **(Table 2, Figure 2E**).

**Figure 2:**
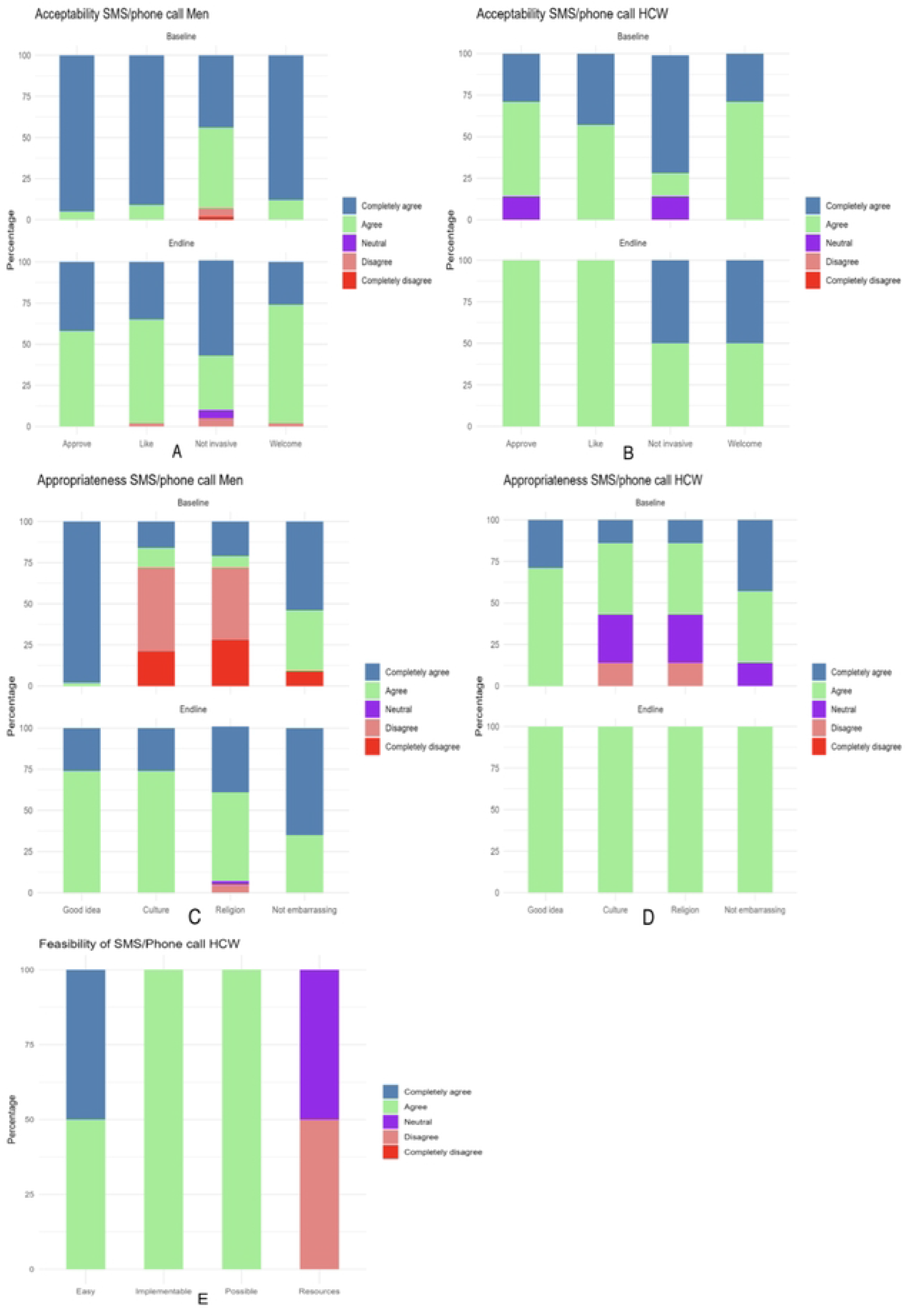
Distribution of Likert scale scores for acceptability, feasibility and appropriateness of SMS/phone call tracing among men and HCWs at Bwaila STI Clinic at baseline and end-line.

### Acceptability, Feasibility and Appropriateness of transport reimbursement among men and HCWs

Overall, transport reimbursements were highly acceptable among men and HCWs at baseline and end-line with most participants completely agreeing and agreeing to the acceptability statements (**Figures 3A and 3B).** Transport reimbursements were appropriate among HCWs at baseline and end-line however, most men completely disagreed or disagreed that transport reimbursement for VMMC were a good fit to their culture and religion (median = 3, IQR: 2, 5). This perception improved at end-line after exposure to the intervention (median = 4, IQR: 4, 5) (p=0.01) (**Table 2, Figures 3C and 3D**). Transport reimbursements were deemed fairly feasible as some HCWs disagreed that they were easy to implement and that resources were available (median = 4, IQR: 2, 4) (**Figure 3E**).

**Figure 3:**
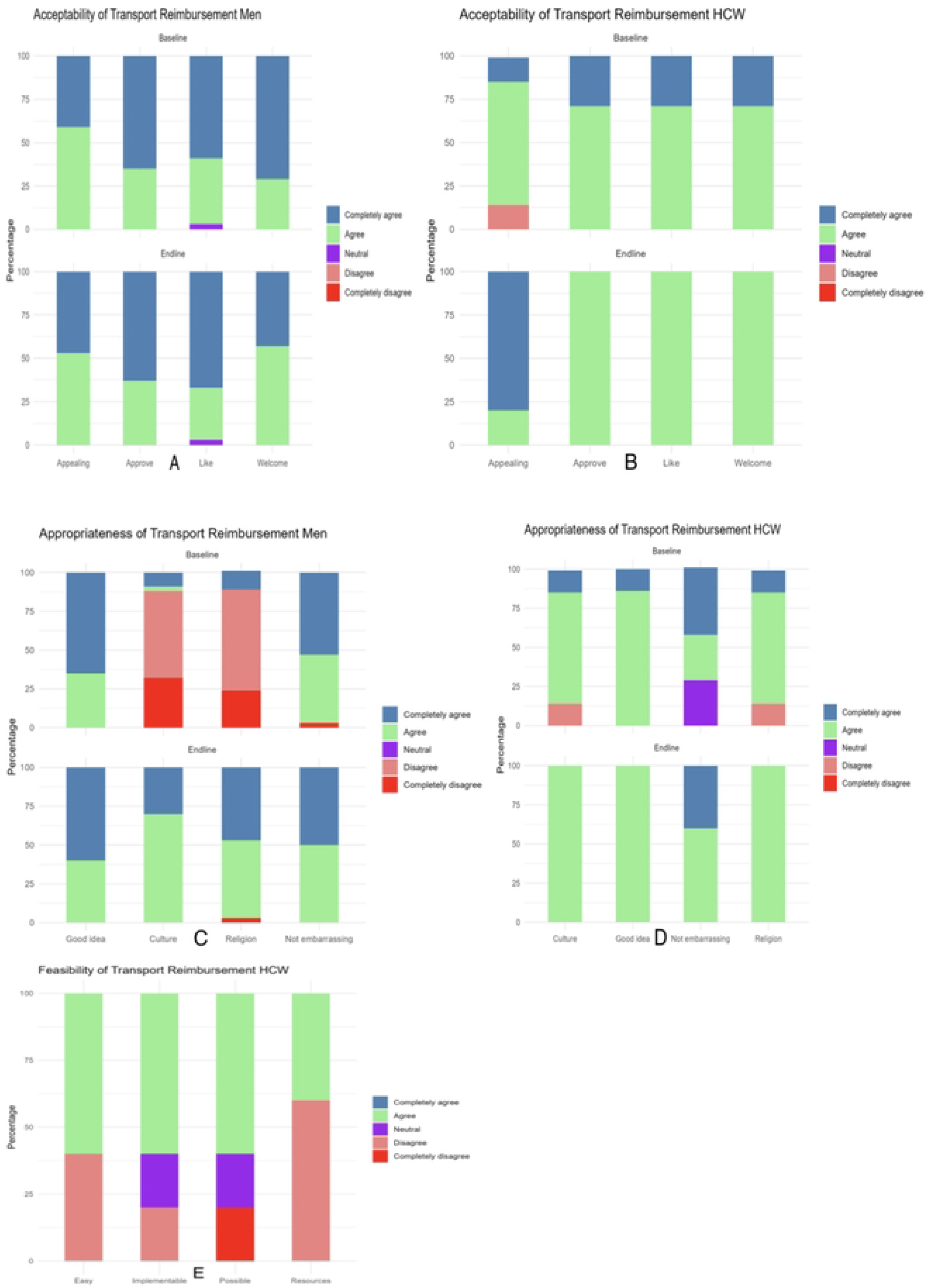
Distribution of Likert scale scores for acceptability, feasibility and appropriateness of transport reimbursement among men and HCWs at Bwaila STI Clinic at baseline and end-line.

### Qualitative

Sixteen (16) uncircumcised men participated in the baseline FGDs and 11 (4 circumcised men and 7 uncircumcised) men in the end-line FGDs. The demographic characteristics were similar to those who participated in the surveys, with a mean age of 27 years (SD ±6) at baseline 28 years (SD ±7) at end-line, and most being married (69% baseline, 45% end-line) and with up to secondary school education (100% baseline, 91% end-line). There were 10

HCWs who participated in the baseline IDIs and seven were interviewed again at the end-line. Similar to the survey participants, most were female nurses with about 10 years of work experience (Table 3).

**Table 3:** Demographic characteristics of healthcare workers who participated in in-depth interviews at Bwaila STI clinic.

|  | HCWs - IDIs |  |
| --- | --- | --- |
|  | Baseline N=10 | *End-line N=7 |
| **Age (years), mean (±SD)** | 37 (±7) | 39 (±9) |
| **Sex n (%)** |  |  |
| Male | 3 (30.0) | 2 (28.6) |
| Female | 7 (70.0) | 5 (71.4) |
| **Cadre** |  |  |
| Nurse | 5 (50.0) | 4 (57.1) |
| ITS counselor | 4 (40.0) | 2 (28.6) |
| Data clerk | 1 (10.0) | 1 (14.3) |
| **Experience (years), mean (±SD)** | 10 (±6) | 10 (±5) |

### Acceptability of IHE

In line with quantitative results, IDIs and FDGs reinforced the acceptability of IHE. The qualitative inputs provided additional insights on what made IHE acceptable and how it could be improved to reach additional people and dispel myths and misconceptions.

#### Importance of information

Men and HCWs reported that they liked intensified health education because it provided information on VMMC which was lacking in the community. They stated that it is difficult for men to undergo VMMC without the right information. An added benefit of IHE was that once men have the right information, they can spread the information to others in the community.

> *“…most people would not decide to go through with VMMC if they have not been taught. The best way would be to spread the information so that they are first taught.”* (End-line FGD2, Respondent 2)

> *“If you teach a person, they will also teach others and they will come back to the clinic with their friends so that even the friends can get better understanding.”* (End-line FGD2, Respondent 6)

However, both men and HCWs recommended that intensified health education should not only be provided at the health facility, but also in the community to extend reach.

> *“It is also important to spread these messages (on VMMC) in the church so that everyone is aware.”* (Baseline FGD2, Respondent 5)

> *“The best way, which would really be helpful is to reach out… you can travel from here to [name of] village and plan for a soccer match between teams, teams that are famous and has players which the people in the rural area have never seen. People will rush to see the players and to watch the football. When the people come, before the match starts, the first thing you can give education.”* (End-line FGD1, Respondent 1)

> *“Some people even go to community leaders and give health talks, they mobilize people in the community. Another way would be to run radio adverts and people who are far can also hear the messages.”* (Baseline IDI, Nurse 4)

#### Dispel myths and misconceptions

Both men and HCWs felt that IHE was a good intervention as it would help to dispel myths and misconceptions about VMMC in the community.

*“This education is very important because it will remove all the misconceptions or things that are against circumcision. As we said, I said that people think that when the foreskin is cut, the government uses it for other purposes. When this education is there, those views that are against the true aims of circumcision will be corrected because people will know and realize that the aim of circumcision is to protect their lives.”* (Baseline FGD2, Respondent 2)

### Appropriateness of IHE

#### Culture and religion

In contrast to the survey results, where concerns were raised by men about appropriateness, there was a general sense from FGDs and IDIs that although culture and religion previously played a significant role in influencing decisions around VMMC, these are now losing their significance. According to HCWs, circumcision is now freely discussed everywhere including churches, hence IHE did not conflict with culture or religion.

> *“I think that the cultures are losing their significance. I think it* (IHE) *would have no effect on culture and I do not think it would discourage people. That is because people already have knowledge on circumcision and so accepting this will not be very hard unlike the way things were previously.”* (Baseline IDI, ITS counselor)

> *“Currently, they talk of circumcision even in the churches. I think the religion that was difficult was Nyau, but I think that even they take part in circumcision, so I don’t see education on VMMC conflict with religion”* (End-line IDI, Nurse)

> *“….issues of circumcision are now part of our normal lives. People now understand the importance of circumcision in the community including among boys or students. Some cultures like the Lomwes or Chewas and religions like Christians that did not believe in circumcision now before believe in it because of the medical benefits.”* (End-line FGD1, Respondent 4)

### Feasibility of IHE

In agreement with the quantitative data, IDIs reinforced the feasibility of IHE. HCWs, the implementers of the intervention, found IHE easy to implement and adoptable as routine practice.

#### Routine practice

Since group health education was already standard practice at the clinic, most HCWs felt that IHE could easily become routine practice and recommended it.

> *“Yes, it* (IHE) *will fit in. It will be combined into what we already do because things like health education and sensitization talks are given on daily basis. As such, things like the health education would just go into the talks that have been happening all along.”* (End-line IDI, Nurse)

#### Easy implementation

HCWs reported that IHE would be easy to implement within the clinic because adding information on VMMC to the pre-existing talks would not feel like additional workload.

> *“Umm, it (IHE) is easy to do because the one who gives the education is a different person and the people who provide other services are different too and so it is easy to do.”* (End-line IDI, ITS Counselor)

> *“As one of the people who gives health talks at this clinic, I thought adding circumcision to our usual health talks worked well since we were just adding extra content to what we already discuss. The talks on circumcision caused some interesting discussions and I could see that people including both men and women were interested to know more.”* (End-line IDI, Nurse)

### Acceptability of SMS/Telephone tracing

The IDIs and FDGs corroborated the quantitative results and reinforced the acceptability of SMS/telephone tracing providing insights on why men and HCWs found this strategy acceptable including recommendations to use SMS/telephone tracing as an additional mode of communication post-circumcision.

#### Reminders

Both men and HCWs appreciated the SMS/telephone tracing strategy as a good way of reminding men who had appointments for VMMC to report for the procedure. As observed by one HCW:

> *“Some people forgot when they had been given a date and so when we sent the SMS, it was a reminder.”* (End-line IDI, Data clerk)

#### Source of encouragement

Beyond being a reminder, most men perceived SMS/telephone tracing as a source of encouragement to men who were doubting their decision to undergo VMMC.

> *“We stay in different areas and you can agree to go to the clinic but as the day comes, you might start to reconsider your decision of going to the clinic. After you receive the reminder, you can get encouraged and even though you changed your mind, you can still decide to go to the clinic.”* (Baseline FGD1, Respondent 6)

#### Private and confidential tracing

An additional advantage raised by several HCWs is that SMS/telephone tracing offers a private and confidential alternative to physical tracing.

> *“Apart from the cost, I think that it is a private way. Some people do not want tracing cars to reach their homes or for the people doing the tracing to go to their homes. On that one, I feel you are maintaining the confidentiality of the potential client who will come for circumcision rather than physically visiting them.”* (Baseline IDI, Nurse)

#### Additional mode of communication

As a recommendation, some respondent suggested that SMS/telephone tracing could be used as a means of communication with the doctor:

> *“It (SMS/Telephone tracing) is a very good thing because after you have been circumcised and you are facing a certain challenge, it can be easy to get in touch with the doctor who sent the message and tell him how you are feeling. In that way, it would help the client save money because if the problem is small, he will easily get assisted.”* (Baseline FGD2, Respondent 7)

> *“Even though we have been educated, we still hear things in the communities and among those things, there may be things that are against what we have learnt, depending on the different cultures as well. This can also give us an advantage of asking the doctors for clarity if there was anything discouraging you.”* (End-line FGD1, Respondent 4)

### Appropriateness of SMS/telephone tracing

In accordance with the survey results, the qualitative data provided more insight into why SMS/telephone tracing was appropriate including suggestions on how to use SMS to reach more people, and also highlighted reasons why SMS/telephone tracing was inappropriate to culture and religion, particularly at baseline.

#### Good idea

Participants reported that one SMS from a HCW has the potential to reach many people. *“The messages are a good idea because they can be used to reach out to many people at once.”* (Baseline FGD2, Respondent 5)

One participant suggested that SMS can be used to spread information on VMMC, and the messages can be shared among people in the community to reach more people.

> *“…..You can spread information on circumcision via phones and we can equally share the messages with other friends and if they want to come, they can come for circumcision.”* (Baseline FGD1, Respondent 5)

#### Culture and Religion

There was a general sense from the FGDs that due to prevailing societal beliefs around promotional messages for various interventions, SMS/telephone reminders would lead to misconceptions in the community. One participant likened the strategy to a previous ‘blood sucking’ misconception around medical outreach services that were mistaken for a ‘cult’ or ‘satanism.’

> *“There are wrong perceptions that we have in our cultures as well. If you receive a message and you tell people that ‘I have received an SMS reminding me to go for circumcision’, people will call it satanic with the times that we are in. The benefits are there, but the negatives are also there because most people believe that receiving an SMS that is encouraging you to do something that involves bleeding or body parts is satanic, that they want you to be a Satanist as well, just like the blood suckers.”* (Baseline FGD2, Respondent 3)

In line with the quantitative data, the qualitative data also demonstrated improved perceptions towards SMS/telephone tracing at end-line as portrayed below:

> *“For me, it was a very good thing to send me a message. I did not have any problems with receiving a message to remind me about circumcision. It did not go against my culture or beliefs or religion. It was just fine.”* (End-line FGD1_Respondent 1)

### Feasibility of SMS/telephone tracing

In accordance with the quantitative data, the IDIs confirmed that SMS/telephone tracing was a straightforward strategy to implement and adopt as routine practice. In addition, the IDIs provided more insight into resource concerns in the quantitative data.

#### Routine practice

Some men were of the view that SMS/telephone tracing was already routine practice because healthcare personnel send numerous messages on different health issues.

> *“Yes, in short, I can explain that as health personnel, there are a lot of messages that you send out. The same way you send messages on other issues, you should do the same with this one* (VMMC) *regardless of how you think people might react.”* (Baseline FGD2, Respondent 2)

This was echoed by *HCWs* who felt that SMS/telephone tracing was not a new thing, comparing this to what happens in the HIV program.

> *“It* (SMS/Telephone tracing) *is also very good because in other cases messages are sent to people to remind them to come and collect ARV’s* (antiretroviral medication) *for instance.”* (End-line IDI, Nurse)

#### Technical and resource challenges

Concerns were raised that SMS/telephone tracing may not be easy to utilize due to technical and resource challenges such as low literacy, lack of phone ownership, and service provider and electricity challenges.

> *“Just to add, I think the SMS would not really be helpful. That is because when we take a person from the rural areas, they use phones yes but all they know is to take calls*. *Even if there is a message, it is hard for them to read it.”* (Baseline FGD2, Respondent 1)

> *“Most people either don’t have phones or they just sold it. It could be that the phone has been damaged and others do not have electricity in their homes. You would find that the day the message is sent, the phone is off.”* (End-line IDI, Nurse)

### Acceptability of transport reimbursement

#### Offset costs

Both men and HCWs felt that transport reimbursement was a good intervention as it would help to offset the cost of transportation.

> *“That is quite a good thought. The reason is that like I said, we are not doing VMMC here. As such, someone might want to undergo it (VMMC) but they might fail because they do not have transport to get to the clinic or to go back home after. That will therefore help other people who might have not had any transport money to come to the clinic for VMMC. That is quite a good thought.” (RIT_012)*

#### Ease transportation

Both men and HCWs alluded to the fact that transport reimbursement enabled men to travel to and from the clinic with ease, especially after undergoing circumcision.

> *“It is good because with a wound, it was hard to travel and as I said, since we are not supposed to engage in hard labor after circumcision, so it is good.” (FGD3_RITE_R4)*

> “*Some people who came to the clinic lived far. So, the money was helpful for them travel back home with ease.” FGD4_RITE_R1*

However, some men thought it would have been better if a car was provided to take men home after circumcision instead of transport reimbursement.

> *“I think it was good for you to provide transport money, it was good. However, I feel it would have been better if you refunded the money used for transport when coming to the clinic but when going back home, you should have dropped us off. I am saying that because some of us live very far. After circumcision I got on a bus. From where the bus dropped me off, I had to get on a bicycle taxi in order to get home. By the time I got home I was in pain. So, it is better if they give you the money you spent when coming to the clinic, but they should drop you off at home afterwards…” (FGD1_RITE_R4)*

#### Attractive incentive

There was a consensus of opinion that transport reimbursement was an attractive incentive as it involved money. Men being breadwinners were more attracted to the money as it would be ‘passive income’ that they can use to cover expenses during the post-circumcision period when they can’t be involved in ‘hard’ work.

> *“The amount of $10 that you have mentioned, they would not only use it for transport. They can also use it for their other needs. Some say that ‘after I am circumcised, what will I eat in the week that I am nursing my wound’. Some people therefore would use it during that one week for their needs while they have the wound so that as the wound heals, they will have the strength to go back to looking for food. So, most people look at it from that angle and not just as transport.” (End-line IDI, Respondent 5)*

However, despite the consensus that transport reimbursement was attractive, some men felt it wasn’t enough, reinforcing the idea that the opportunity costs of VMMC go beyond transport.

> *“Yes, just to add, the strategy is good, but we really need to stress on the financial support for the family. We can be reimbursed the transport, but the main problem is how we will sustain our households. The transport reimbursement will only be useful for that same day and yet we will have to stay home for several days. Those other days are the ones that would be problematic.” (FGD1_RITE_R3)*

### Appropriateness of transport reimbursement

#### Culture and religion

There were conflicting reactions concerning the appropriateness of transport reimbursement particularly its fit with culture and religion. Most men felt transport reimbursement would not be well received by Muslims:

> *“For us Muslims, we would really be concerned and our leaders specifically. That is because it would completely reduce the number of people being circumcised in our culture. Most people would rush to be circumcised at the hospitals because as we have already said, money is a problem. So, there would be disappointment among our leaders because of the money. Few people would go to them compared to those who would go to the hospitals.” (Baseline FGD2, Respondent 4)*

In contrast, most HCWs felt the strategy was a good fit.

> *“I think that the cultures do not have much influence. I think a lot of people would be happy to receive money for transportation regardless of their culture or religion.” (RIT_001)*

#### Result in misconceptions

Both men and HCWs raised concerns that transport reimbursement would be viewed as coercive, feeding into misconceptions in the community.

> *“People would have different views. Some would think it is good while others would say that ‘because you have been circumcised, you have sold your foreskin’. They would think that the government has come up with this system as a way of buying your foreskin. (Baseline IDI, Counsellor)*

### Feasibility of transport reimbursement

#### Unsustainable

It was a unanimous opinion by HCWs that transport reimbursement would not be made routine as there would be no financial resource for the government to implement this, as reflected in the following quote.

> *“So, this strategy is good because we will reach the people and help them. But the issue is after the study is over and we want to continue. Sustainability becomes a challenge…. I do not think that government would be giving transport”. (End-line IDI,* Nurse)

## DISCUSSION

Among a population of men and healthcare workers at an urban STI clinic, the RITe intervention was generally acceptable, feasible and appropriate. Participants expressed that all strategies were acceptable. However, the cultural and religious appropriateness of some strategies were questioned in some cases, particularly at baseline. Notably, appropriateness improved at end-line for SMS/telephone tracing and transport reimbursement but not for IHE. From the HCW perspective, intensified health education was perceived to be the most feasible strategy in contrast to SMS/telephone tracing and transport reimbursement.

Intensified health education was rated as highly acceptable among men and HCWs at baseline and end-line. The high acceptability was corroborated in the qualitative results where most men and HCWs perceived IHE as a source of important information and a means of dispelling myths and misconceptions on VMMC. This was further evidenced by men and HCWs advocating for IHE to be extended to the community. IHE was deemed highly feasible resulting in HCWs recommending IHE to be integrated into routine health education at the clinic. Health education/awareness continues to be one of the most effective, feasible and acceptable interventions for improving uptake of VMMC in most settings [19,20]. Several approaches to health education have all been acceptable in VMMC programs. These include sensitizing community or religious leaders/organization [21–23], schools [22], influencers [21,24], mass media [23] and peer recruiters and women [21,22,25]. Other approaches are using satisfied clients [26], mobilizers [22,23,26], one-on-one counselling [26], house-to-house sensitization [26], distribution of educational materials [23,26], and targeting men hangouts [26].

Although some participants in our study postulated that culture and religion no longer play a significant role in influencing peoples’ perceptions on VMMC and that talking about circumcision in the community is normal, this perspective was not shared universally. For example, culture and religion still influenced IHE appropriateness level in the surveys and religious concerns were flagged in relation to transport reimbursement during FGDs. Another observation is that appropriateness of IHE was conflated with appropriateness of circumcision in general as men did not distinctly separate the two. We believe this contributed to the views on appropriateness in the surveys and FGDs. In a recent systematic review of barriers to uptake of VMMC in priority countries, negative perceptions of VMMC due to culture and/or religion was the most cited barrier [7]. Our findings agree with the suggestions that VMMC communication should separate circumcision from cultural or religious identities and emphasize the medical benefits of circumcision for all men; community and religious leaders should be engaged in VMMC awareness efforts in order to shift norms [7].

Our findings also indicated high acceptance of the SMS/telephone tracing strategy among men and HCWs at baseline and end-line. Participants liked, approved and welcomed the strategy as a reminder for VMMC appointments. Men specifically liked SMS/telephone tracing as a source of encouragement to undergo VMMC and as an opportunity for communicating with HCWs in case of questions or challenges during the recovery period. HCWs liked the strategy as a means of maintaining private and confidential communication with VMMC clients. An SMS intervention was found acceptable by providers and men in another study where SMS was used for post-circumcision follow up [27]. The messages reminded clients of the signs of poor healing and provided a platform to communicate with providers which reduced unnecessary in-person clinic visits,[27] both reasons being similar to those elicited in our study.

Participants raised a few concerns on appropriateness of SMS/telephone tracing at baseline. However, their perceptions of the appropriateness of SMS/telephone tracing improved at the end-line. This was reflected in the FGDs where men feared that SMS/telephone tracing would results in misconceptions in the community. One participant reported that messages on VMMC would be perceived as ‘satanic’ in the community as they would be likened to messages that people receive enticing them to ‘cults’ and religion. However, these concerns did not reappear at end-line, after exposure to the intervention. In contrast, an SMS intervention for VMMC was equally culturally appropriate in two recent studies from Zimbabwe [27] and South Africa [28] that assessed acceptability and appropriateness of SMS in post-operative follow-up among VMMC clients.

The feasibility of SMS/telephone tracing was hampered by lack of mobile phone access, low literacy and technical constraints such as network coverage. These limitations are not uncommon in resource-limited settings. In a systematic review on barriers to use of mobile health in improving health outcomes in developing countries, in which 60% of the studies were SMS-based, some of the most common barriers were lack of access to phones and inability to use them [29]. In contrast, studies have reported high feasibility and usability of SMS interventions; however, they included cell phone ownership as part of the eligibility criteria [27,28]. Therefore, it is not surprising that feasibility and usability was high in these studies because of the correlation between frequency of phone usage and knowledge of the SMS technology [29], which is higher among people who are literate and own phones.

In Malawi, the Global Systems for Mobile Communication Association estimated that access to mobile phones was around 57.2% in 2023 [30]. In our study, about 61% of men owned a phone, while 30% of those without a phone provided an alternative phone number belonging to someone they trusted and 9% did not own a phone. It is likely that the low cell phone coverage in the country compounded by the technical barriers may have influenced feasibility assessment among HCWs. Nonetheless, SMS is very popular in developing countries, and it can be used for interventions and health education [29].

Transport reimbursement attracted a lot of attention among men in this study. The strategy was rated highly acceptable by men and HCWs. Our findings agree with regional results that monetary incentives are generally attractive and effective [19,20]. In this study, transport reimbursement was viewed as a way of offsetting the cost of commuting to health facilities and easing travel especially post-circumcision, and a source of encouragement to undergo VMMC. However, some men preferred receiving transport reimbursement for travel to the facility only and being ferried home in a vehicle post-circumcision to reduce chances of being in pain before arriving home.

Though considered attractive, other men reported that reimbursing transport was not adequate. They preferred receiving additional support to feed their families during the recovery period as financial concerns due to missing work or losing income and family survival during the recovery period are common concerns among men in resource-constrained setting [31]. In contrast, some men perceived the transport reimbursement as more than adequate and that it would be coercive as men would undergo circumcision because of the money [20].

Interestingly, the appropriateness of transport reimbursement attracted mixed reactions. On the one hand, some men reported that transport reimbursement would conflict with the Muslim religion as most people would prefer VMMC due to the monetary incentive over ‘ritual’ circumcision, which is done at a cost to the client. On the other hand, HCWs reported no conflict with religion. Apart from religion, some reported that transport reimbursement would reinforce misconceptions in the community, that people would think the government is buying foreskins for trade with western countries. This misconception has also been reported in other priority countries [32].

Healthcare workers unanimously viewed transport reimbursement as an unsustainable strategy as most governments in low- to mid-income countries would not be able to provide monetary incentives as routine practice. Even though financial incentives are the most effective strategy for increasing demand for VMMC, sustainability is indeed a concern in developing countries [33]. Hence, it is generally recommended that financial incentives should not be considered at a primary demand-creation strategy but as an additional strategy among many tools [33]. Financial incentives should be considered when a certain threshold of coverage has been reached to encourage late adopters to undergo circumcision, thereby improving the cost-effectiveness [33].

The strength of this study is in the extensive assessment of acceptability, feasibility and appropriateness through a mixed-methods approach which allowed us to triangulate findings from both sources to better explain our results. On the contrary, our study may have limited generalizability in rural settings. We hypothesize that barriers such as culture and religion are likely to be more dominant in rural settings than in urban settings [9]. However, considering that most of men in the study had up to secondary school education, findings could be somewhat similar if this study was repeated in a rural setting. In addition, the strategies were likely more acceptable, appropriate and feasible in an STI clinic setting as men with STIs could have seen a greater value in undergoing VMMC as part of treatment of their STI compared to outpatient clinic settings where VMMC is offered/provided in most facilities.

Our intervention with intensified health education, SMS/telephone tracing and transport reimbursement strategies was found acceptable, feasible and appropriate by men and healthcare workers as strategies for increasing demand for VMMC at an urban STI clinic. Among the strategies, intensified health education was highly acceptable, feasible and appropriate. More awareness is required to address cultural and religious barriers to VMMC. SMS interventions are gaining popularity and are bound to have more impact as countries continue to develop. Monetary incentives such as transport reimbursement are highly attractive but they should not be considered as a primary demand-creation strategy.

## Data Availability

Data cannot be shared publicly because of the country ethics and institutional policy. Data are available from the UNC Project Malawi Institutional Data Access (contact via) for researchers who meet the criteria for access to confidential data.

## Acknowledgements

We would like to thank the Lilongwe District Health Office, the Bwaila STI clinic team, Mrs. Chimwemwe Kana and the VMMC team from Jhpiego Malawi Office and VMMC program coordinators from Lumbadzi Health Center, Kang’ona Health Center, Chitedze Health Center, Area 25 Health Center and Area 30 Health Center for their support in conducting this study. We also thank the men who participated in this study.

This project was supported by NIH Fogarty grant # D43 TW009340, D43 TW010060 and D43 TW009774-06. The study sponsors have not had any role in the study design and will not play a role in collection, management, analysis and interpretation of data; writing the report; and the decision to submit the report for publication.

## Author Contributions

MM, MCH, SJ and CC contributed to the conceptualization and design of the study. MT, JT, BN, NB, EM, EJ, DY, BK, and MK assisted with data collection and curation. MM, EK and JC assisted with statistical analysis. MT, JT, MM and SJ contributed to the qualitative analysis. MM drafted this manuscript. All authors reviewed and approved the manuscript.

